# The Effects of India’s COVID-19 Lockdown on Critical Non-COVID Health Care and Outcomes: Evidence from a Retrospective Cohort Analysis of Dialysis Patients

**DOI:** 10.1101/2020.09.19.20196915

**Authors:** Radhika Jain, Pascaline Dupas

## Abstract

India’s COVID-19 lockdown, one of the most severe in the world, is widely believed to have disrupted critical non-COVID health services. However, linking these disruptions to effects on health outcomes has been difficult due to the lack of reliable, up-to-date health outcomes data. We identified all dialysis patients under a statewide health insurance program in Rajasthan, India, and conducted surveys to examine the effects of the lockdown on care access, morbidity, and mortality. 63% of patients experienced a disruption to their care. Transport barriers, hospital service disruptions, and difficulty obtaining medicines were the most common causes. We compared monthly mortality in the four months after the lockdown with pre-lockdown mortality trends, as well as with mortality trends for a similar cohort in the previous year. Mortality in May 2020, after a month of exposure to the lockdown, was 1.70 percentage points or 64% (p=0.01) higher than in March 2020 and total excess mortality between April and July was estimated to be 22%. Morbidity, hospitalization, and mortality between May and July were strongly positively associated with lockdown-related disruptions to care, providing further evidence that the uptick in mortality was driven by the lockdown. Females, socioeconomically disadvantaged groups, and patients living far from the health system faced worse outcomes. The results highlight the unintended consequences of the lockdown on critical, life-saving non-COVID health services that must be taken into account in the implementation of future policy efforts to control the spread of pandemics.

## INTRODUCTION

On 24 March 2020, the Government of India ordered one of the most stringent nationwide COVID-19 lockdowns in the world to control virus spread (Hale et al. 2020). The lockdown was announced with four hours’ notice, barred people from leaving their homes, required non-essential commercial establishments and transport services to close, and was enforced strictly with penalty of arrest (Ministry of Home Affairs 2020). National restrictions were eased 10 weeks later, at the beginning of June, but localized restrictions continued in areas with high case counts.

Although critical health services were officially exempt from the lockdown, the media reported widespread disruptions to routine and emergency non-COVID care due to transport and curfew barriers for patients and health workers, hospitals turning patients away, and supply chain disruptions that affected medicine access and costs (Indian Express 2020, IndiaSpend 2020, New York Times 2020). Medical services under government health insurance programs across the country decreased by 51% and the national Health Management Information System (HMIS), which collates monthly reports from the public health system, reported dramatic decreases in preventive and curative care (IndiaSpend 2020, Smith et al. 2020).

A second wave of COVID-19 infections engulfed India in 2021, with the country reaching close to 400,000 new cases daily in early May 2021, far exceeding the first in magnitude (Johns Hopkins 2021). In part, due to the perceived costs of the first lockdown, the government eschewed nationwide lockdowns in favor of localized curfews and closures in COVID-19 hotspots. Nevertheless, the health system has been overwhelmed by the surge, though the economic costs have been more muted (The Lancet 2021, Indian Express 2021).

Lockdowns can help “flatten the curve” of new infections, buying the health system time to prepare for and respond to the pandemic (Figueiredo et al. 2020, Flaxman et al. 2020, Islam et al. 2020). Recent research suggests India’s 2020 lockdown reduced the growth in COVID-19 incidence, largely by restricting mobility (Winter et al. 2021). However, the indirect costs of restricting mobility and economic activity may be sizeable (Douglas et al 2020, Haug et al 2020). In India, these costs were exacerbated by the hasty imposition of the lockdown, which gave the health system and households little time to prepare (The Lancet 2020, Cash and Patel 2020). People with chronic health conditions are particularly vulnerable to pandemic-related care disruptions (Modesti et al. 2020). Largescale, quantitative vidence of disruptions to non-COVID health care during the 2020 lockdown is now emerging from around the country, particularly for chronic conditions like cancer, kidney disease, diabetes, and tuberculosis, but their health impacts have not been measured (Jain et al. 2021, Cilloni et al. 2020, Kusuma et al. 2021, Ranganathan et al. 2021, Singh et al. 2021). Careful measurement of the indirect morbidity and mortality effects of the lockdown is critical to understanding the full consequences of the pandemic and how to prepare health systems better for future disease outbreaks (Beaney et al. 2020, Kiang et al 2020, Setel et al 2020).

Quantifying the impacts of such disruptions on morbidity and mortality has been difficult in the Indian context due to the unavailability of reliable and high frequency data measuring health outcomes and cause-specific mortality. Because the COVID-19 lockdown may have reduced deaths from some causes, such as road accidents, evaluating its effects requires disaggregated cause-specific mortality. The vast majority of deaths in India occur at home, rather than at health facilities, are not included in the national Civil Registration System, and have no certified cause of death (Jha et al. 2005). While government health insurance programs collect real-time data on hospital services provided, they record no details on patient morbidity or mortality. The HMIS data are typically of low quality, underrepresent care in the private sector, and do not provide complete morbidity or mortality outcomes (Sharma et al. 2016). The Sample Registration System, which estimates age-specific death rates through population surveys, does not provide details on cause of death and the most recent report only provides total mortality estimates through 2018 (SRS Bulletin).

Given the dearth of reliable, updated, and publicly available data on health outcomes in India, evaluating the effect of the COVID-19 lockdown on non-elective health services requires the identification of patients in need of such care and the collection of primary data. We used insurance claims filed under a largescale government health insurance program to identify patients requiring dialysis, a form of non-elective chronic care, during the lockdown. We conducted phone surveys with their households to estimate the effects of the lockdown on health care access, morbidity, and mortality in the four months following its imposition. The goal of this study was not to quantify the trade-off between averted COVID-19 mortality and the non-COVID-19 costs of the lockdown, but, more simply, to document the extent to which the lockdown worsened morbidity and mortality for chronic care patients. To our knowledge, we provide some of the first quantitative measures of the effects of India’s 2020 COVID-19 lockdown on excess mortality from a non-COVID health condition.

## DIALYSIS IN INDIA

We focused on dialysis, a form of life-sustaining long-term hospital care for patients with end stage chronic kidney disease (CKD). CKD is the 17^th^ leading cause of deaths globally and 8th in India (GBD 2015). Although access to dialysis treatment is relatively low, the expansion of government health insurance has increased its reach and a 2019 study estimated that there are approximately 175,000 CKD patients on dialysis in India (Jha et al. 2019). Dialysis removes waste, salts, and excess water to prevent their build up in the body; regulates levels of potassium, sodium, and bicarbonate in the blood; and controls blood pressure. The typical patient requires three- to four-hour sessions, two to four times each week for the duration of their life or until they get a kidney transplant. Disruptions to dialysis treatment result in the accumulation of fluids and toxins in the body, and can cause extreme swelling, nausea and vomiting, difficulty in breathing and urinating, and other symptoms. Missing dialysis visits or shortening their duration is associated with large increases in hospitalization and mortality (Anderson et al. 2009, Abdel-Kadar et al. 2009, Saran et al. 2003, Zoraster et al. 2007). Recent work documents substantial disruptions to dialysis care in India during the lockdown, but does not link them to health outcomes (Ramachandran and Jha 2020, Prasad et al. 2020, Smith et al. 2020).

## METHODS

### Study Population and Data

We conducted a retrospective cohort analysis to study care-seeking and health outcomes before and after the COVID-19 lockdown. The study population was all dialysis patients enrolled in a government health insurance program that covers the poorest two-thirds of Rajasthan’s population. The Ayushman Bharat Mahatma Gandhi Rajasthan Swasthya Bima Yojana (AB-MGRSBY), is a statewide government health insurance program in Rajasthan, India. It entitles approximately 50 million low-income individuals to free secondary and tertiary care at over 1200 empaneled hospitals. Household eligibility is based on state poverty lists. All members are automatically enrolled and face no premium, deductible, or copay. Hospitals file claims in real-time for every patient visit through the government’s electronic filing system. These claims data, which include patient name, phone number, address, hospital visited, and services received, provide one of the only ways of directly identifying patients utilizing hospital care in Rajasthan.

We obtained access to all administrative claims data filed under the program from its launch in 2015 through October 2019. Using the last 3 months of these data, we identified all patients on dialysis under insurance between August and October 2019. Because dialysis is long-term and non-elective care, patients on dialysis in late 2019 would continue to require it through the COVID-19 lockdown if still alive. Therefore, these patients provide an ideal group to study the effects of the lockdown on critical chronic care.

To collect data on health care access and outcomes through the lockdown period, we conducted phone surveys using patient contact numbers in the administrative records. The survey was conducted with the patient or the person in the household most knowledgeable of their care if the patient was unwell or dead, and collected data on dialysis visits in the month prior to the lockdown, disruptions to care due to the lockdown, morbidity, hospitalization, the date and cause of death, and basic demographic and socioeconomic characteristics. We also embedded open-ended questions into the surveys to collect qualitative descriptions from patients about care disruptions faced. We completed the first round of surveys between late May 2020 and mid-June 2020, and follow-up surveys in July and August with all patients alive at the time of the first survey to track complete mortality through July. To help put survey results into context, we also conducted structured interviews with 15 hospitals from 10 of Rajasthan’s 33 districts.

### Outcomes and Measures

We measured disruptions to dialysis care by asking whether patients faced each of the following problems during the lockdown: their dialysis hospital was closed; it was open but refused to provide them services; they could not travel to their hospital due to lack of transport or curfews; they had to switch to a different hospital from their primary dialysis hospital; their hospital increased charges over the typical payment; they had difficulties obtaining their dialysis medicines. Reported dialysis visits were used to create an indicator for any decrease in visits between the month before and after the lockdown. We created an individual care disruptions index using the first component of a principal-components analysis (PCA) of all of these indicators, and standardized it over the study sample for ease of interpretation.

To track changes in monthly mortality over the months before and after the lockdown, we created a mortality time series between December 2019 and July 2020. We determined the month of death for all dead patients by cross-checking two measures - the exact month of death and the number of months that had lapsed since the death at the time of survey - as well as the cause and location of death to ensure it was not unrelated to dialysis. Monthly mortality, or the likelihood of death, was calculated as the number of deaths each month as a share of people alive at the beginning of that month.

Morbidity and hospitalization were reported for the four weeks prior to the survey, in each round of the survey. A morbidity index was constructed from a PCA of indicators for whether the patient experienced the following symptoms that are known to follow disruptions to dialysis care and can be reported by patients: swelling of the face, hands, legs, or body; vomiting or nausea; extreme tiredness or weakness; difficulty breathing; difficulty urinating; and muscle cramps. The index was standardized over the sample. Hospitalization was an indicator for any in-patient hospital visit.

To examine heterogeneity in outcomes by socioeconomic and demographic characteristics, we created binary classifications for female, age under 45 years, lower caste group (scheduled caste or tribe), and low assets. Patients were classified as low asset if they had a below median score on an asset index created from a PCA of a list of assets they own, such as a motorcycle, television, or air-conditioner. Additionally, we geocoded all dialysis hospitals in the AB-MGRSBY program as well as the residence locations for 88% of surveyed patients using addresses in the administrative data that were verified by survey. We calculated the distance from patient residence to the district administrative headquarters, the largest city in the district, as a proxy for remoteness, and to the closest dialysis hospital as a proxy for distance from the health system. For heterogeneity analysis, we created binary classifications for above and below the median distance in the study sample, which was 45km for distance to the city and 35km for distance to a dialysis hospital.

### Comparison Cohort

Because the COVID-19 lockdown was implemented across all of India at the same time, there are no comparable “untreated” populations in 2020. Therefore, our analysis primarily focused on changes in mortality trends within our study cohort over the four months before and after the lockdown was imposed. In addition, we constructed a historical comparison cohort to allow us to account for potential seasonal trends or monthly fluctuations in mortality and to examine whether 2020 mortality follows historical trends. Using the same administrative insurance claims data, we identified all patients on dialysis under insurance between August and October 2018. Because phone surveys with these patients would likely suffer from attrition and recall bias, we instead used the share of patients that permanently dropped out from dialysis care each month to proxy for the share of people that died in that month. We have previously validated this measure of mortality through surveys and confirmed that 80% of patients that dropped out of claims had died and the remaining 20% had dropped out for reasons besides death (exiting the insurance program, out of state migration, kidney transplant, or discontinuation of treatment for financial reasons). Monthly mortality in the historical cohort was, therefore, calculated as the 80% of the share of all patients on dialysis each month that dropped out of treatment in that month.

### Statistical Analysis

To examine changes in monthly mortality before and after the lockdown, we estimated a non-parametric discrete-time logistic model with the probability of death as the outcome and binary indicators for each month from December 2019 to July 2020. We excluded October and November 2019 from the analysis because survey dropout was likely to be highest in these months and could bias mortality estimates, but complete death counts for the full period are provided in the supplement table S3. Adjusted models included indicators for age above 45 years, female sex, lower caste group, and low assets, and continuous measures of months on dialysis and total dialysis visits at baseline. To examine historical mortality trends, we ran the same unadjusted model on the comparison cohort with indicators for each month from December 2018 to July 2019, as well as a model adjusted for age above 45 years and total dialysis visits prior to enrollment. Standard errors were clustered at the patient level in all models to account for autocorrelation.

To analyze the association between lockdown-related care disruptions and health outcomes, we restricted the sample to patients alive at the end of April (and, therefore, exposed to at least one month of the lockdown), and estimated linear OLS regressions, with heteroskedasticity-robust standard errors, of morbidity, hospitalization, and death between May and July on the care disruptions index. Adjusted models included indicators for age above 45 years, female sex, low caste group, and low assets, as well as continuous measures of months on dialysis and total dialysis visits at baseline. Sensitivity to using logistic regression models was also assessed.

We also examined whether the lockdown had differential effects on care-seeking and outcomes by sex, age (above 45 years), lower caste, and low assets. First, we estimated bivariate and multivariate logistic regressions of an indicator for having experienced any disruption on binary indicators for each subgroup characteristic. Second, we calculated the percentage point change in monthly mortality between March and May for each subgroup from logistic regressions controlling for all other subgroup characteristics and dialysis history. Lastly, we estimated the association between care disruptions and health outcomes separately for each subgroup.

## RESULTS

We identified 3,183 patients on dialysis under insurance between August and October 2019 across Rajasthan. Of these, 2,234 (70.2%) had a reachable phone number, and 94% of them consented to participating, resulting in a study sample of 2,110 patients (supplementary Table S1). Attrition may be due to numbers being entered incorrectly in the insurance data, households changing numbers, or unused numbers being deactivated and reassigned to new households. Of the 1,392 patients alive at the time of the first survey, 1,177 (85%) completed a follow-up survey. Successfully surveyed patients were disproportionately male (69%), had a mean age of 46 years, and had been on dialysis for a year, with 5 visits per month on average, when they were enrolled into the study (Table 1). Surveyed patients were almost identical in these characteristics to patients not reached, but had been on dialysis for slightly longer, increasing our confidence that attrition did not meaningfully bias our sample (supplement Table S2). On average, patients lived 25km from the nearest hospital providing dialysis care. The vast majority of patients (83%) were visiting a private hospital for their dialysis treatments.

**Table 1:** Study sample characteristics. Dialysis history is drawn from administrative claims data on all dialysis treatment received prior to enrollment in the study in October 2019. Distance to district HQ is the distance in kilometers to the administrative center of the district, a proxy for distance to the closest city. Distance to dialysis hospital is the distance to the closest hospital providing dialysis under insurance, a proxy for distance from the health system.

|  | Mean | SD |
| --- | --- | --- |
| Age (years) | 46.0 | 14.7 |
| Female (%) | 31.1 |  |
| Scheduled caste/tribe (%) | 18.7 |  |
| Education (years) | 11.3 | 2.6 |
| Distance to district HQ (km) | 48.8 | 38.8 |
| Distance to dialysis hospital (km) | 19.7 | 17.7 |
| <u>Dialysis history at baseline</u> |  |  |
| Private dialysis hospital | 83.2 |  |
| Monthly dialysis visits | 5.0 | 3.5 |
| Months on dialysis | 12.6 | 11.5 |
| Observations | 2,110 |  |

### Disruptions to Dialysis Care

Over 63% of patients reported a disruption in access to dialysis care during the lockdown (Figure 1). 42% of patients reported being unable to reach their hospital due to travel barriers. Open-ended interviews indicated that travel barriers were largely due to difficulties finding transport and obtaining official exemptions from district administrative and health authorities to travel to the hospital. 15% of patients found the hospital was closed or refused to provide care, 11% faced increased hospital charges, and 23% had to switch to a different hospital from the one they typically visit. Interviews suggest supply chains disruptions reduced medicine availability and increased prices. As a result, 17% of patients could not obtain their necessary medicines. Patients faced a 172% increase in payments per visit, driven largely by increased charges at private hospitals. 22.2% of patients experienced a decline in monthly dialysis visits between March and April and the average decline was 6%. Hospital interviews confirmed patient reports, but also suggested that hospitals faced their own constraints. Seven of the 15 hospitals reported having to close during the lockdown due to staffing and supply shortages. Those that continued operating reported a substantial drop in patient visits, which they credited to transportation barriers, and larger hospitals reported receiving patients displaced from nearby hospitals that had closed.

**Figure 1:**
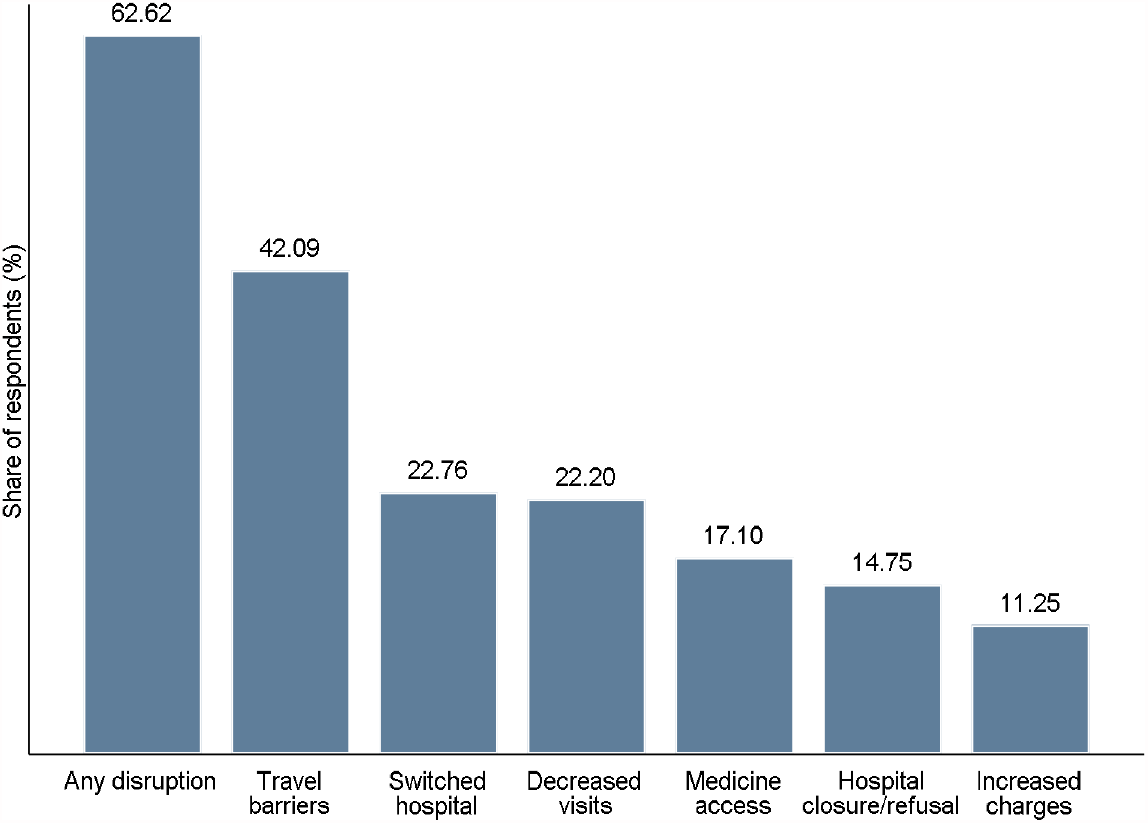
Disruptions to dialysis care during the COVID-19 lockdown. The figure presents the share of patients that reported experiencing disruptions to their dialysis care due to the lockdown between imposition of the lockdown in March 2020 and the survey conducted in May-June 2020.

### Association Between Care Disruptions and Outcomes

Among patients alive at the end of April and exposed to at least one month of the lockdown, a 1SD increase in in the morbidity index (p<0.001), 3.1pp increase in the probability of hospitalization (p=0.002), and 2.1pp increase in the probability of death (p=0.013) in the period from May to July 2020 (Table 2). These effects are sizeable relative to an overall 14.2% hospitalization and 10.8% mortality hazard over those months. Controlling for sociodemographic characteristics and dialysis history did not change these relationships. They also remain robust to using logistic instead of OLS regression models for the binary hospitalization and death outcomes (supplemental Table S5).

**Table 2:** Association between lockdown-related care disruptions and health outcomes. The table presents linear regressions of health outcomes between May and July 2020 on a standardized index of lockdown-related disruptions to dialysis care. The sample is patients alive at the end of April and, therefore, exposed to at least one full month of the lockdown (N=1,489). Morbidity is a standardized index of symptoms known to follow disruptions to dialysis. Hospitalization and death are binary outcomes. Covariates in adjusted regression models include age, sex, caste group, month of dialysis initiation, and lifetime dialysis visits at baseline.

| Outcome | Association Between Care Disruptions Index and Health Outcomes |  |  |  |  |  |
| --- | --- | --- | --- | --- | --- | --- |
|  | Unadjusted |  |  | Adjusted |  |  |
|  | Estimate | 95% CI | P | Estimate | 95% CI | P |
| Morbidity Index | 0.172 | 0.125 - 0.219 | <0.001 | 0.171 | 0.123 - 0.218 | <0.001 |
| Hospitalization | 0.031 | 0.012 - 0.051 | 0.002 | 0.030 | 0.010 - 0.050 | 0.003 |
| Death | 0.021 | 0.004 - 0.038 | 0.013 | 0.025 | 0.008 - 0.042 | 0.004 |

### Changes in Mortality Trends

Monthly mortality declined steadily from December 2019 to March 2020 (Figure 2), due to early deaths among the most vulnerable patients, such as the elderly and lower caste (supplement Figure S1). Mortality in May 2020, after a full month of exposure to the lockdown, increased sharply to 4.37%, a 1.70pp or 63.60% (p=0.01) change relative to mortality in March, prior to the lockdown. Controlling for sociodemographic characteristics and patient dialysis history increased this difference to 1.85pp or 67.77% (supplement Figure S2). Mortality declined in June and July 2020, but never dropped below March levels, indicating that deaths in May were not simply a displacement of mortality from the subsequent two months. Because we did not reach all households in the follow-up survey, true mortality in this period may be higher than measured. Only four patients who died in June and July were reported testing positive for COVID-19. Excluding them reduces mortality to 3.16% in June and 2.69% in July.

**Figure 2:**
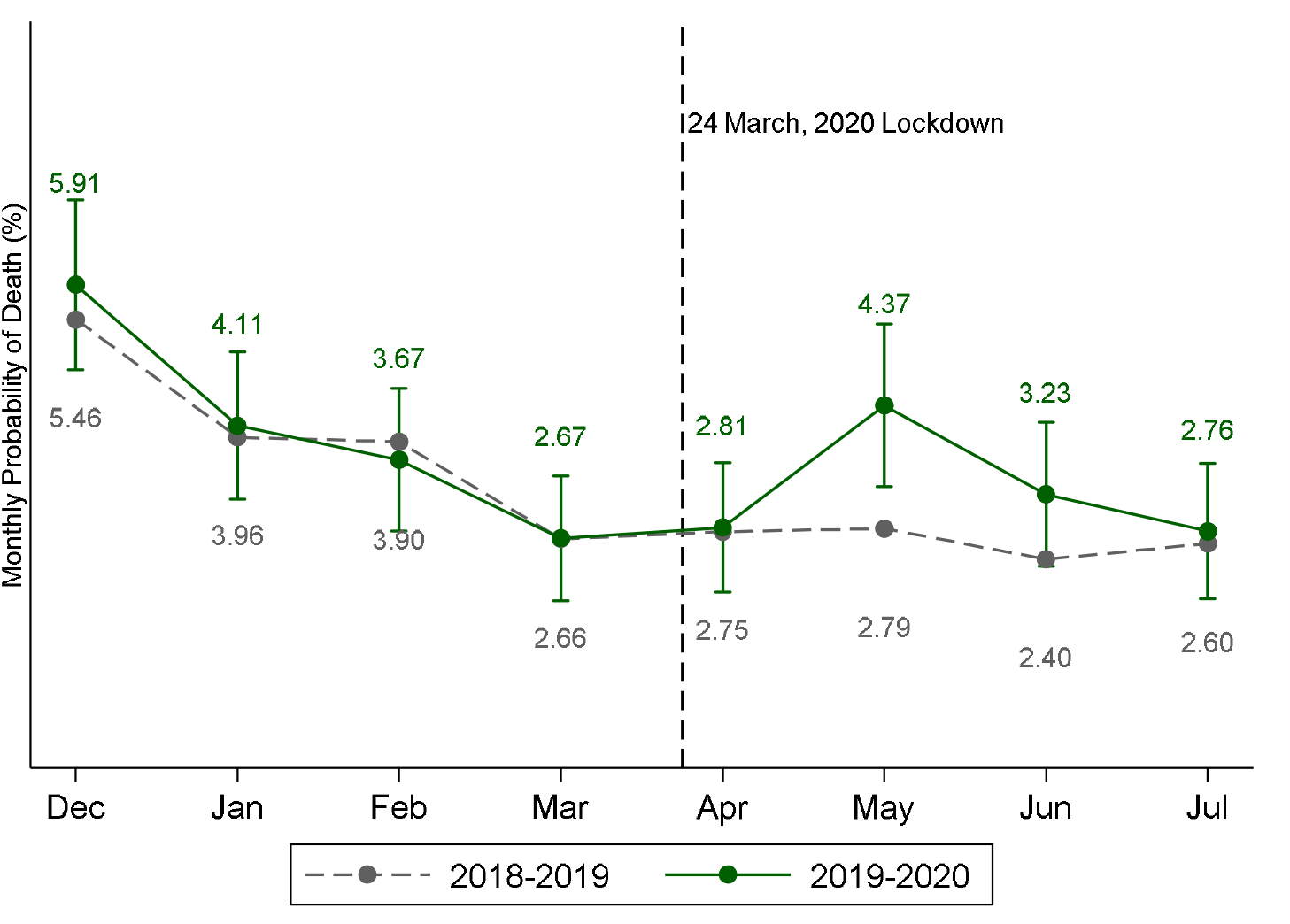
Unadjusted monthly dialysis mortality in the surveyed and historical cohorts. The solid line presents monthly likelihood of death, or the share of people still alive that die in each month, for the surveyed cohort, from an unadjusted discrete time logistic regression model with indicators for each month from December 2019 through July 2020. Vertical bars represent 95% confidence intervals. The dashed line presents the monthly hazard for the comparison cohort of patients on dialysis in August-October 2018 from a similar model with indicators for each month from December 2018 through July 2019. Mortality estimates have been converted into percentage terms for ease of interpretation. Covariate-adjusted models in the online supplement look almost identical.

We compare this to mortality over the same months in the previous year in the historical comparison cohort. The historical cohort followed a statistically similar trend between December 2018 and April 2019, but exhibited no increase in May 2019 or the subsequent months. Comparison of trends in the study and historical cohorts suggests that the sharp increase in May 2020 is not explained by monthly or seasonal fluctuations and that, absent the lockdown, monthly mortality would have remained similar to or slightly below March levels in 2020, as it did in 2019.

To estimate total excess mortality in the four months following the COVID-19 lockdown, we calculated the difference between observed mortality between April and July 2020 and what mortality would have been if the March 2020 mortality probability of 2.67% had applied each month. Out of 1,532 patients alive at the end of March, 192 had died by the end of July (12.54% mortality), whereas 157 would have died had the March rate (10.25% mortality), resulting in 22.3% total estimated excess mortality in the four months following the lockdown (supplement Table S4). This is likely to be a conservative estimate, as both pre-lockdown trends and the historical comparison suggest that mortality would have continued to decline in the absence of the lockdown rather than remain at March 2020 levels.

### Subgroup Heterogeneity in Lockdown Effects

Although the lockdown was universal, its effects on care-seeking were worse for vulnerable and remote households. Being lower caste, poorer, and living further away from a city or a dialysis hospital had large and significant positive associations with the likelihood of facing any care disruption (Table 3). Poverty had the strongest relationship and remained significant in multivariate regressions controlling for other socioeconomic characteristics and dialysis history.

**Table 3.** Association between patient characteristics and likelihood of facing care disruptions. The table presents estimates from logistic regressions of an indicator for “faced any lockdown-related care disruption” on indicators for patient characteristics. The multivariate model includes all patient characteristics from the bivariate models.

|  | Outcome: Faced Any Care Disruption |  |  |  |  |  |
| --- | --- | --- | --- | --- | --- | --- |
|  | Bivariate |  |  | Multivariate |  |  |
|  | OR | 95% CI | P | OR | 95% CI | P |
| Female | 0.974 | 0.780-1.216 | 0.815 | 0.979 | 0.768-1.247 | 0.862 |
| 45yrs or younger | 1.143 | 0.932-1.402 | 0.200 | 1.063 | 0.850-1.330 | 0.593 |
| Lower caste | 1.333 | 1.003-1.773 | 0.048 | 1.152 | 0.849-1.563 | 0.363 |
| Low assets | 1.618 | 1.315-1.990 | <0.001 | 1.495 | 1.191-1.876 | 0.001 |
| Far from city | 1.302 | 1.038-1.635 | 0.023 | 1.212 | 0.962-1.528 | 0.103 |
| Far from hospital | 1.360 | 0.995-1.861 | 0.054 | 1.269 | 0.923-1.744 | 0.142 |

Figure 3 presents the change in mortality in May, relative to March, by subgroup. Underlying data are presented in supplemental Table S6. The increase in mortality was largest and significant for females (3.56pp, p=0.022), patients 45 years or younger (2.94pp, p=0.006), poorer patients (2.81pp, p=0.020), those living further from a city (3.51pp, p=0.013) and those living further from a dialysis hospital (5.64pp, p=0.031). The associations between the care disruptions index and morbidity, hospitalization, and death were also stronger for these subgroups (supplement Figure S3). The greater disruptions to care faced by poorer and remote patients noted above could partly explain their worse outcomes, but we find no evidence that females experienced greater disruptions. An alternate explanation for high female mortality may be lower care-seeking by households, consistent with prior literature on gender bias in health care in India (Anonymous 2021, Kapoor et al. 2019, Shaikh et al. 2018). Females had fewer monthly dialysis visits at baseline (4.8 visits) relative to males (5.1 visits, p=0.062), which could contribute to worse health prior to the lockdown. Additionally, care disruptions were associated with increased morbidity for both males and females, but with increased hospitalization only for males, and larger increases in mortality for females, suggesting that households may have been less likely to seek hospital care for females that faced complications (supplement Figure S3).

**Figure 3:**
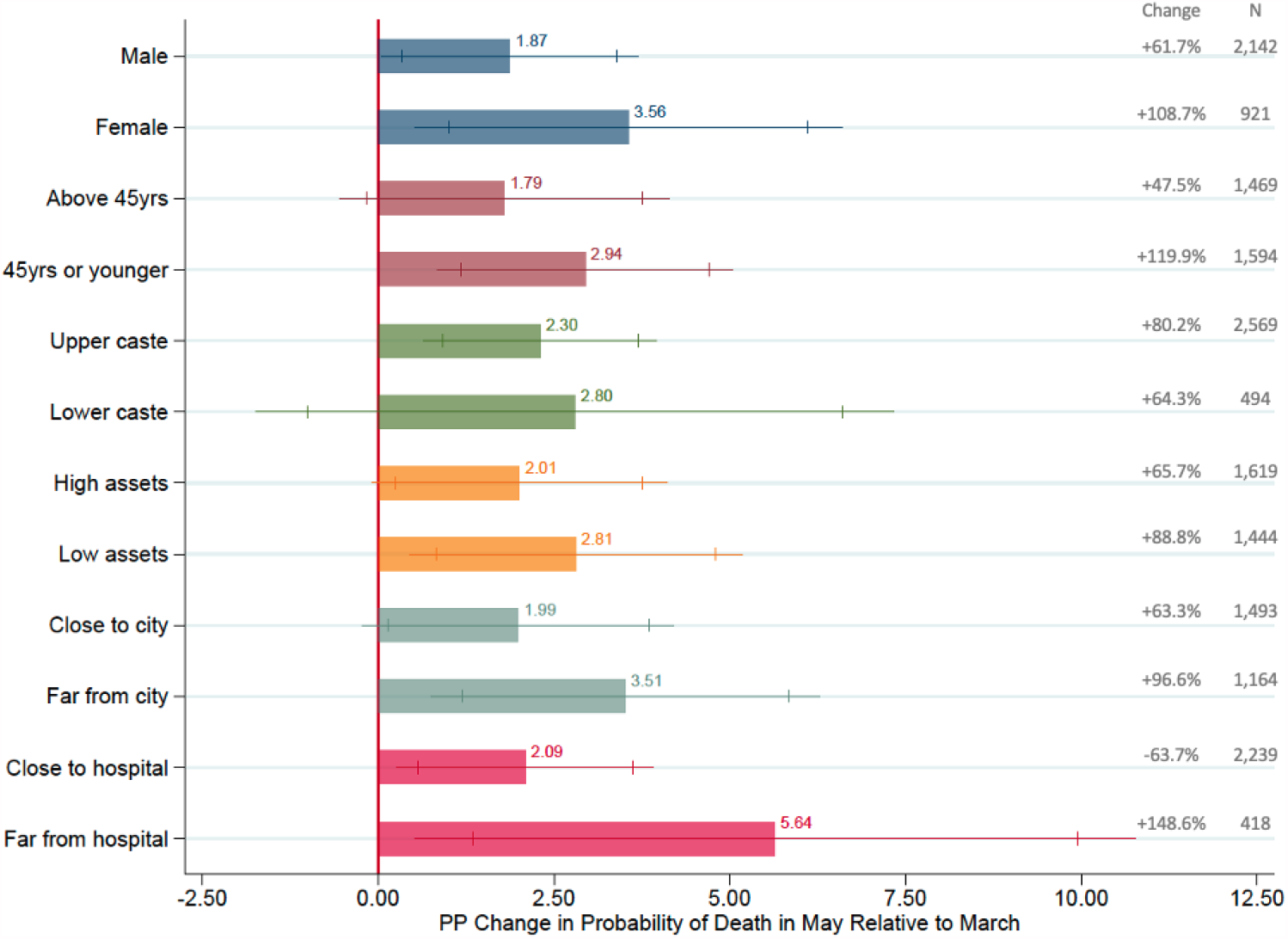
Changes in mortality between March and May by subgroup. Bars represent the percentage point change in mortality between March and May 2020 for each subgroup. Each model is adjusted for all other characteristics (indicators for age, sex, caste, and asset group), as well as total dialysis visits prior to enrollment. All underlying numbers are in supplement Table S6. Lines represent 90% and 95% confidence intervals. The grey figures on the right present the increase between March and May as a percentage of March mortality and the number of observations within each subgroup. Because the figure represents the change in probability of death, the observations are the sum of persons alive in March (1,574) and in May (1,489).

## DISCUSSION

We studied the indirect effects of India’s COVID-19 lockdown on critical, chronic, non-COVID care and outcomes among a large cohort of low-income dialysis patients. Two-thirds of patients faced disruptions to their care during the lockdown. Travel barriers, hospital closures, service refusals, and difficulties obtaining medicines were common causes. As a result, 23% of patients switched away from their primary hospital, 22% experienced a decrease in monthly dialysis visits, and 11% faced increased treatment charges. Lockdown-related disruptions to dialysis care were significantly positively associated with morbidity, hospitalization, and mortality in the months just after its imposition. After declining steadily from December, mortality in May 2020, after a month of exposure to the lockdown, increased sharply and was 1.70pp (63.60%) higher than in March 2020. Mortality for a similar cohort over the same months in the previous year, proxied by permanent dropout from dialysis care, followed a steady downward trend between December and July. The timing and size of the increase in mortality in May 2020 relative to pre-lockdown trends in 2020, as well as to trends in estimated mortality in the previous year, strongly suggest that it was due to the nationwide COVID-19 lockdown. Estimated overall excess mortality in the four months after imposition of the lockdown was 22%.

The excess mortality we measure is unlikely to be due to COVID-19 infections. Although patients on dialysis are at higher risk of COVID-19 infection and complications and excess mortality among dialysis patients infected with COVID-19 in India has been documented, if this were driving mortality in our setting, we would expect deaths to increase over time as the virus spread and to affect older patients more (Gansevoort and Hilbrands 2020, Kakkanattu et al. 2021). However, the increase in mortality was largest in May, soon after the lockdown and before the virus had spread widely, and was greater among patients under age 45 than among older patients who have higher COVID-19 mortality risk. Therefore, we believe our estimates are capturing the indirect effects of the pandemic through lockdown related disruptions to care. Nevertheless, given that COVID-19 testing rates in India were relatively low at the time of the study, we cannot rule out the possibility that it contributed to the deaths we measured.

Importantly, we find that the effects of the lockdown on care access and mortality were more severe among patients of low socioeconomic status, and those in remote locations underserved by the health system. These results indicate that a universal lockdown policy is likely to have larger adverse effects on already vulnerable subpopulations, consistent with the literature on the unequal distribution of both direct and indirect effects of the pandemic (Bambra et al. 2020, Cash and Patel 2020, Marmot and Allen 2020). Women also experience a larger increase in mortality than males do (though the comparison is not statistically significant), which may be because they receive worse care. It is critical that future pandemic control efforts take these distributional consequences into account and put in place extra protections for populations that have limited financial and geographic access to health services.

Our study population is all dialysis patients in the AB-MGRSBY government health insurance program. Given that the poorest half of Rajasthan’s population is enrolled in AB-MGRSBY, our study is representative of close to all low-income dialysis patients in the state. While our analysis is restricted to one type of critical chronic care in one state, our findings are likely to be indicative of the serious but largely undocumented health effects of severe disruptions to dialysis and a range of similar critical chronic care services in other states in India. A 2019 study estimated that there are approximately 175,000 patients on chronic dialysis across India and recent research suggests that a large share of these patients suffered severe disruptions to their care, similar to those we document (Jha et al. 2019, Prasad et al. 2020, Ramachandran and Jha 2020). A nationwide analysis of dialysis patients under government health insurance programs targeting low-income households, a population very comparable to the one we study, finds a 6% reduction in dialysis visits, very similar to our results (Smith et al. 2020). One study of COVID-19 mortality among dialysis patients also finds evidence of increased mortality among COVID-19-negative patients, which may be attributable care disruptions very similar to those reported in this study (Kakkanattu et al. 2021). Evidence is also emerging of disruptions to critical care for other conditions, such as cancers, cardiovascular emergencies, and TB in India (Cilloni et al. 2020, Kusuma et al. 2021, Ranganathan et al. 2021, Singh et al. 2021). For example, the same analysis of government health insurance programs finds a 64% decline in oncology care and an 80% decrease in critical cardiovascular surgeries (Smith et al. 2020). While the precise magnitude of health effects of care disruptions may vary by condition, our results suggest they may go undetected without careful research tracking cause-specific morbidity and mortality.

A strength of this study is the use of existing administrative government health insurance claims data to rapidly identify and remotely survey a large sample of poor dialysis patients in need of critical care during the lockdown. A limitation of this approach is the substantial dropout between identified dialysis patients and those reached for survey by phone due to patient phone numbers entered incorrectly in the claims data, households switching phone numbers, and unused numbers being deactivated. However, dropout rates are similar to those in other phone surveys and surveyed patients were statistically similar to those not reached, increasing confidence that attrition did not bias our study sample (Singh et al. 2021). Furthermore, these issues are most likely to affect patients that died soon after October 2019 and cannot explain the increased mortality in May 2020. Another limitation is that our outcomes are based on self-reported data and not clinical measures, which were infeasible due to the pandemic. However, our primary outcome is mortality, which is likely to be reliably reported, and the recall period for all outcomes was relatively short. Finally, as we only measure mortality through July, our estimates may be a lower bound on the full health costs of the lockdown. We found that disruptions to dialysis were lower but persistent in July and August, after the lockdown was eased, suggesting that adverse health effects may have continued to unfold past the study period (supplement Figure S4).

An important outstanding question concerns the extent and health impacts of disruptions caused by the massive second wave of COVID-19 infections that India experienced, with over 400,000 new daily cases in early May 2021 (Johns Hopkins 2021). Although there has been no national lockdown, reports suggest access to non-COVID care has been severely affected because health facilities are heavily focused on COVID-19 care or refuse to provide critical chronic care to COVID-19-positive patients (Aljazeera 2021, The Guardian 2021). Our results from the first wave provide strong reason to think that the second wave will have sizeable impacts on non-COVID morbidity and mortality among patients needing critical care, adding to the heavy toll of the pandemic. It is critical that these effects be carefully measured. Our study demonstrates that phone surveys combined with administrative data from existing government health programs provide a powerful way of measuring health impacts.

Lockdowns can reduce COVID-19 transmission (Figueiredo et al. 2020, Flaxman et al. 2020, Islam et al. 2020, Winter 2021). However, they have substantial indirect health and economic costs that may outweigh their benefits relative to other pandemic control strategies in some contexts (Cash and Patel 2020, Douglas et al. 2020, Haug et al. 2020, Modesti et al. 2020). The tradeoffs are complex and the optimal policy, as well as the full range of costs and benefits, will depend on local conditions (The Lancet COVID-19 Commission Task Force). Our findings highlight the unintended consequences of India’s lockdown on critical non-COVID health services that must be taken into account in the implementation of future policy efforts to control the spread of pandemics.

## Data Availability

Replication code and deidentified survey data will be made available at the time of publication on the author websites.

## Notes

### Competing Interest Statement

The authors have declared no competing interest.

### Funding Statement

This research was funded by a grant from the Bill and Melinda Gates Foundation, awarded through the J-PAL CaTCH Initiative special COVID-19 funding window

### Author Declarations

Stanford University, USA (protocol no. 41683); Institution for Financial Management and Research, India

