## Supplemental Materials for "The Effects of India’s COVID-19 Lockdown on Critical Non-COVID Health Care and Outcomes: Evidence from a Retrospective Cohort Analysis of Dialysis Patients"

Online Supplementary Materials

### **Table S1. Survey success rate**

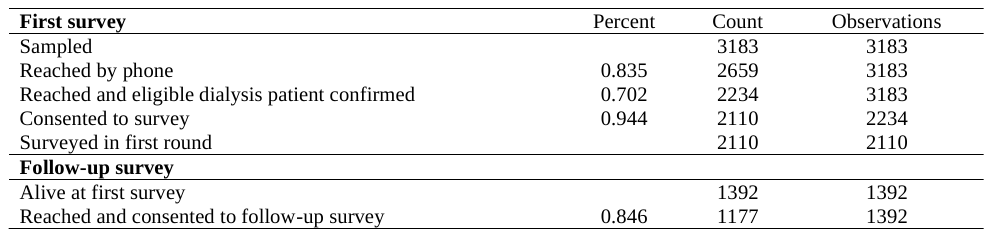

### **Table S2. Baseline characteristics of patients by survey status**

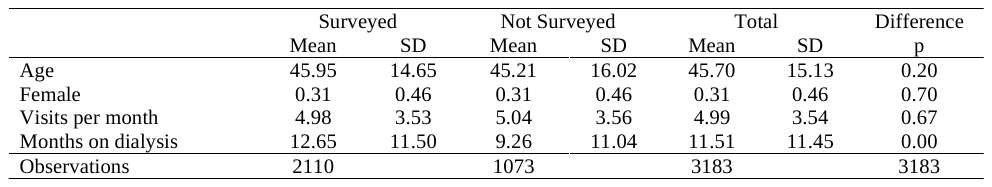

### **Table S3. Complete death counts in survey and comparison cohorts**

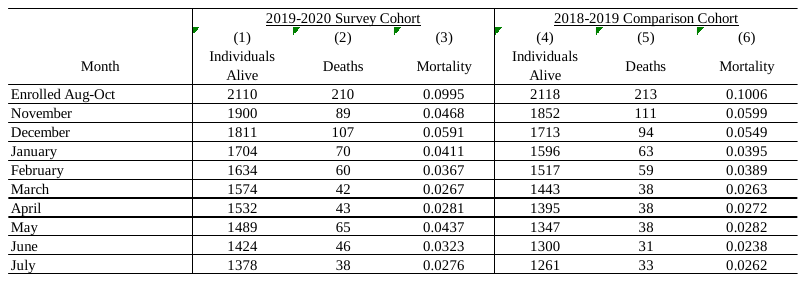

### **Table S4. Excess mortality estimates**

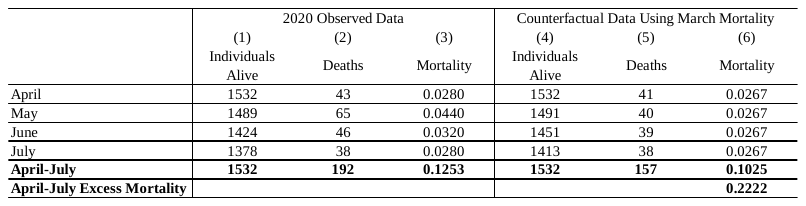

### **Table S5. Association between care disruptions during lockdown and post-lockdown health outcomes: Logistic models**

|  | Hospitalization | | |  | Death | | |
| --- | --- | --- | --- | --- | --- | --- | --- |
|  | OR | 95% CI | P |  | OR | 95% CI | P |
| A. Unadjusted |  |  |  |  |  |  |  |
| Care Disruptions Index | 1.262 | 1.103 - 1.444 | 0.001 |  | 1.222 | 1.053-1.418 | 0.008 |
| B. Adjusted |  |  |  |  |  |  |  |
| Care Disruptions Index | 1.239 | 1.079-1.422 | 0.003 |  | 1.243 | 1.067-1.448 | 0.005 |

The table presents from logistic regressions of the two binary health outcomes on the care disruptions index, corresponding to Table 2 in the paper, which presents OLS estimates.

**Table S6. Changes in mortality between March and May by subgroup**

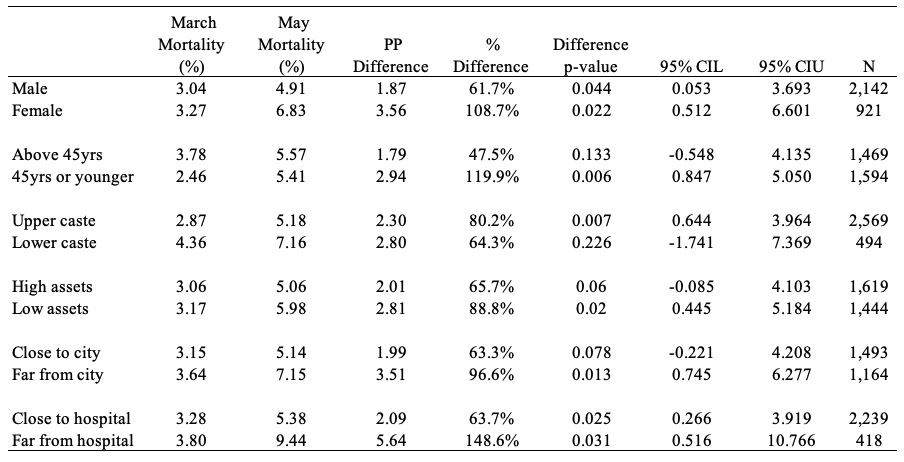

The table presents March and May 2020 mortality by subgroup, the percentage point and percent difference in mortality between the two months, the p-value and 95% confidence intervals for this comparison, and the number of observations within each subgroup. Each model is adjusted for all other characteristics (indicators for age, sex, caste, and asset group), as well as total dialysis visits prior to enrollment. The percentage point difference in mortality in column 3 is also presented graphically in Figure 3 of the paper. Note that the number of observations reflects person-months in March (1574) plus those in May (1489) – see table S3.

### **Figure S1. Unadjusted monthly dialysis mortality by subgroup**

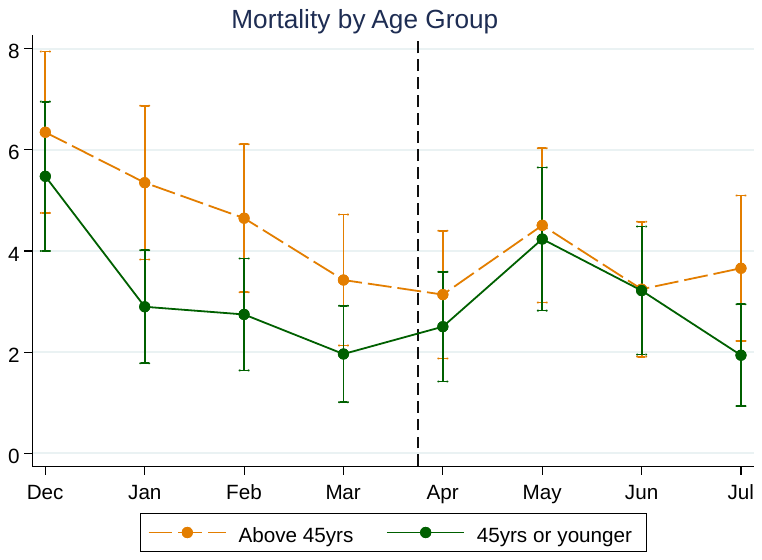

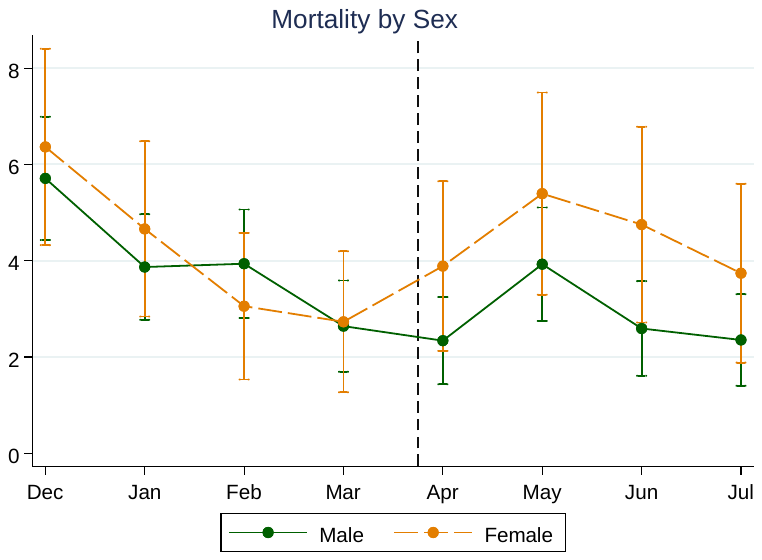

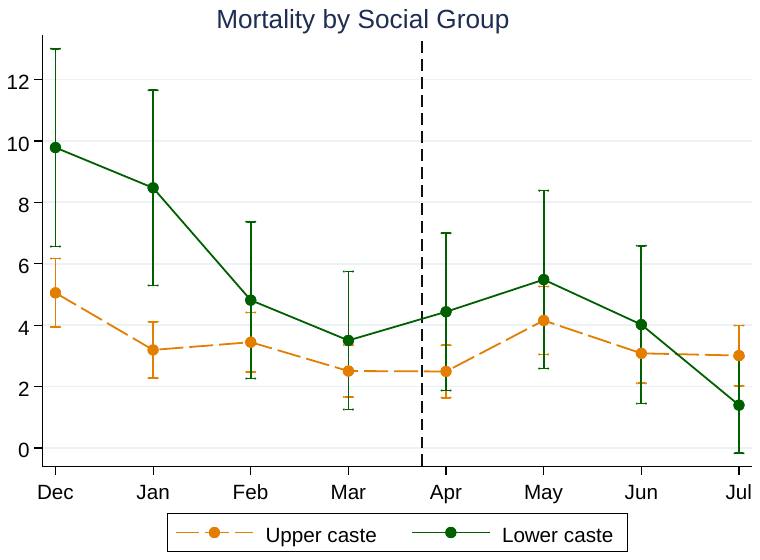

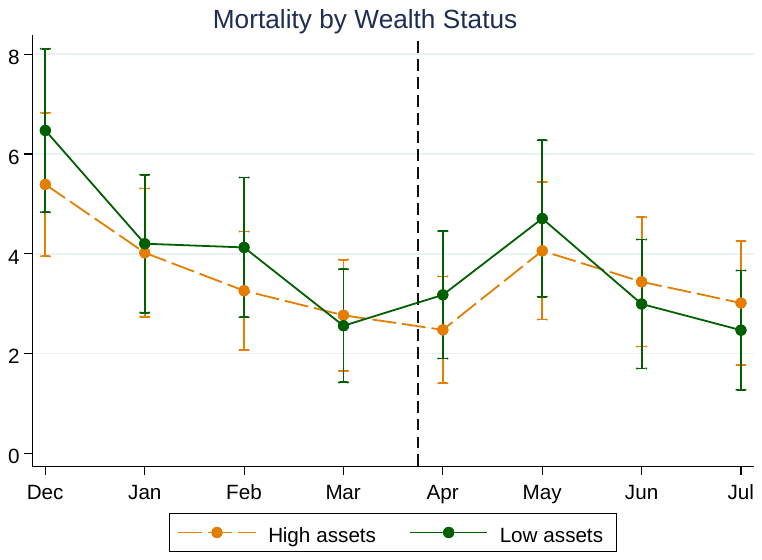

### **Figure S2. Covariate-adjusted monthly dialysis mortality in the surveyed and historical cohorts**

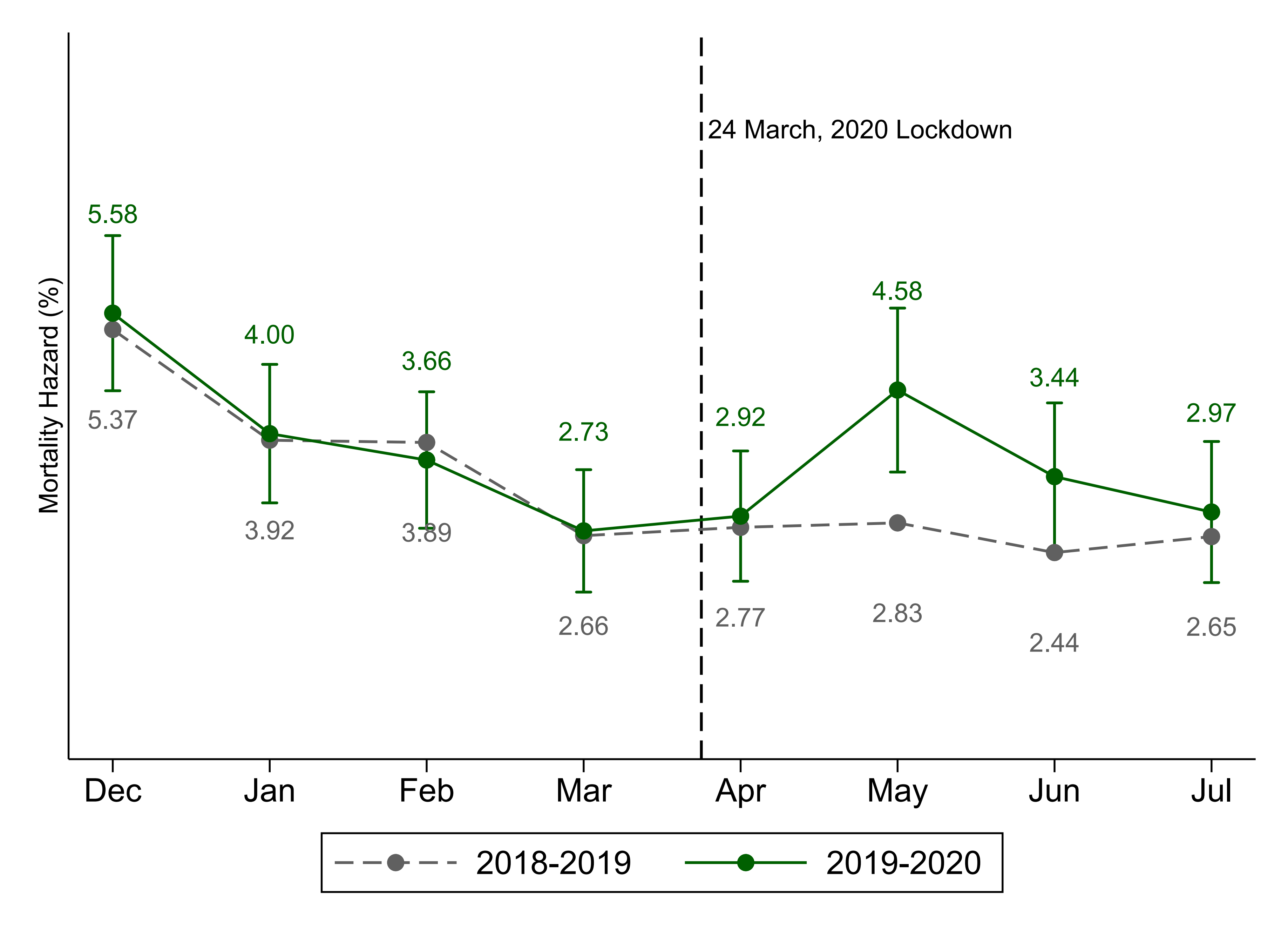

The figure presents corresponds to Figure 2 in the paper, but estimates are adjusted for age, sex, caste group, month on dialysis, and lifetime dialysis visits at baseline for the 2019-2020 cohort and age, sex, months on dialysis, and lifetime dialysis visits at baseline (variables available in the claims data) for the 2018-2019 cohort.

### **Figure S3. Associations between disruptions and health outcomes by subgroup**

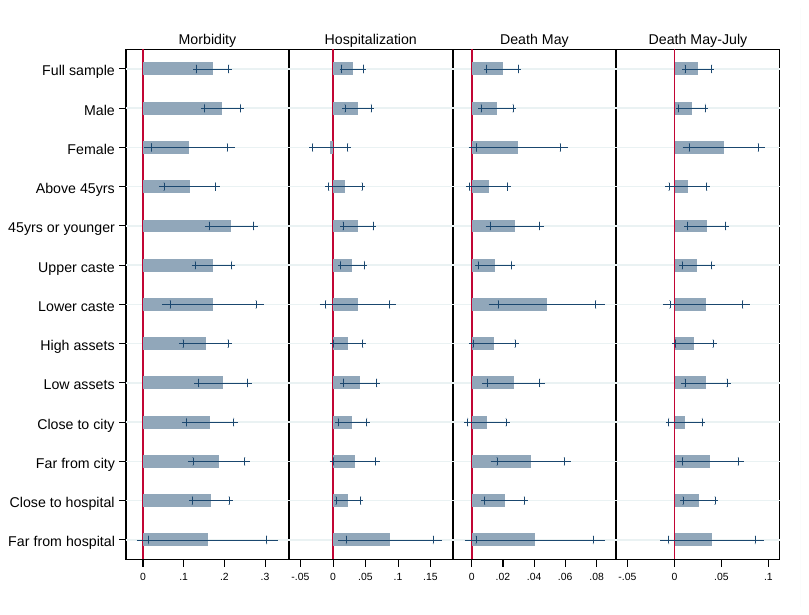

The figure presents associations between the care disruptions index and May-July health outcomes by subgroup for the 1,489 patients alive through the end of April and exposed to at least one month of the lockdown. Each bar is the coefficient on an indicator for the subgroup characteristic from an OLS regression with robust standard errors and controls for all other characteristics (age, sex, caste group, and asset class) and dialysis history at baseline. The outcomes are the standardized morbidity index, and indicators for hospitalization, death in May, and death between May and July.

### **Figure S4. Disruptions to dialysis care in July and August**

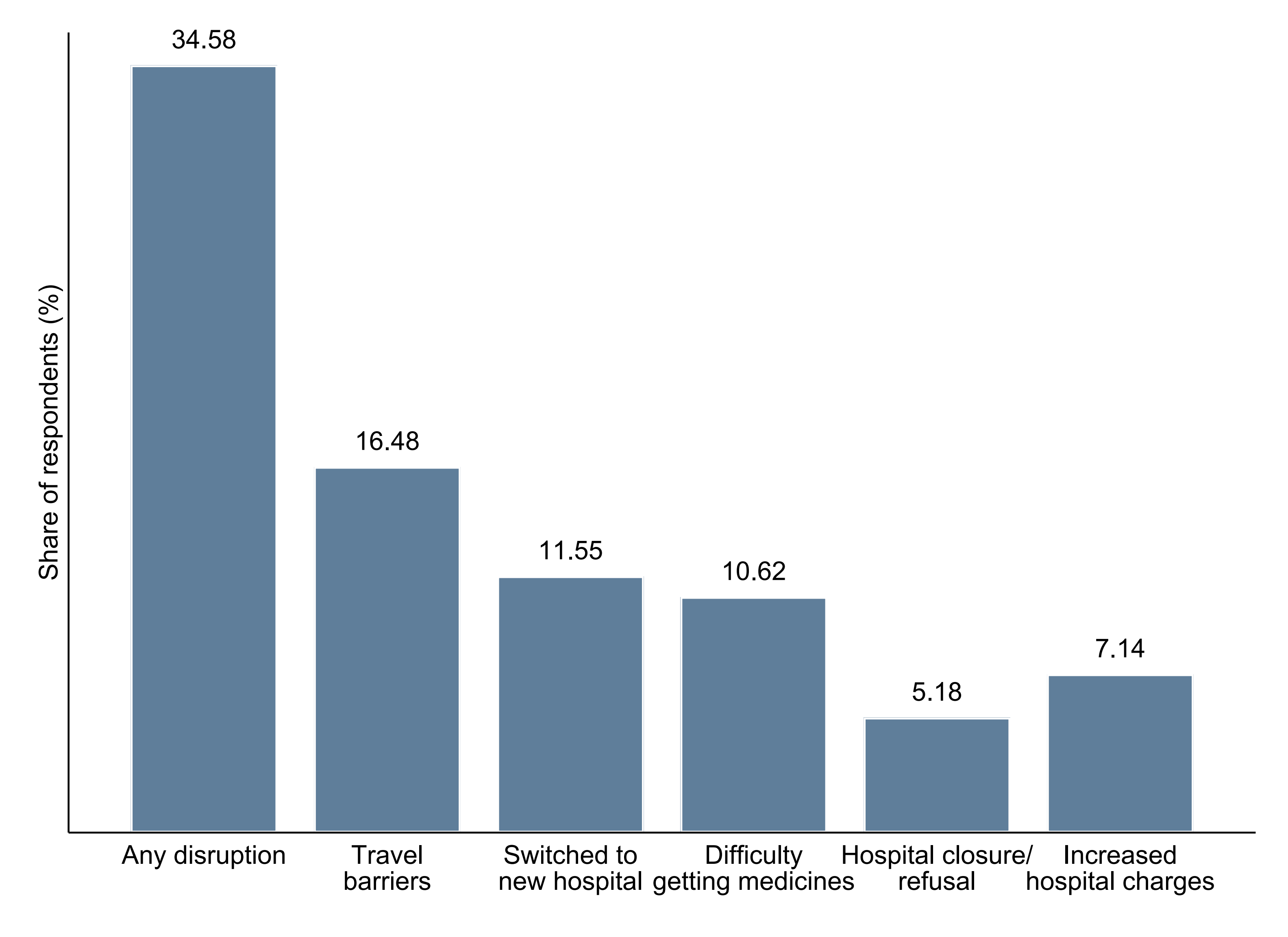

The figure presents the share of people that reported experiencing disruptions in the four weeks prior to the survey, for all patients re-interviewed in July and August. These results suggest that disruptions to dialysis care continued well after the nationwide lockdown was released and may continue to have effects on morbidity and mortality in the coming months.

### **Supplementary Text**

### **Qualitative interviews with patients and hospitals**

To gain a more nuanced understanding of patients’ experiences, we built open-ended questions into the survey. Surveyors were instructed to provide prompts, such as “Have you experienced any other health problems or difficulties in getting dialysis care due to the coronavirus lockdown? Can you describe them to me?” and to encourage respondents to answer freely. The research team listened to voice recordings of these answers and compiled them into structured summaries. The qualitative results confirmed that the list of barriers to care-seeking we included in the survey were comprehensive and covered the key proximate causes of care disruptions: hospital closure or service refusal, increased costs of care, travel/transport barriers, having to switch from their primary hospital to a different one, and difficulty obtaining medicines. They also reveal that many patients experienced a series of different barriers that had compounded effects on their care. Excerpts from some open-ended answers:

*“The hospital was closed on visits and refused treatment as well. […] We did not have a mode of transport to reach the hospital, I had to try very hard to arrange one. […] As a result, I could only get 3 dialysis visits in total instead of 8.”*

*“There was no doctor in the hospital. No one gave us any information where to find one, there was just one ward boy who did not know anything. […] For getting a treatment that my life depends on, I had to get slips from doctors, permissions from District Collector, stand in long queues...”*

*“There is this particular medicine that is required for dialysis procedure and that is not currently available in the market. We have not been able to find it for the last few days. […] Private dialysis centers have increased their rates - it has become unaffordable for us. We don't have money to seek treatment in a private hospital, we have lost our jobs so don't have enough money to go get treated in a private hospital, hence I had to take my wife very far to find a public hospital for dialysis. […]* *Every time we change a hospital, they issue a new set of tests and we have to get them because they will not give us dialysis otherwise. […] Due to shutdown of public transport, I have to now take my wife to dialysis. Because of that neither can I work nor I can be with my wife during her visits.”*

*“I went to two private hospitals, both of them were closed, hence I had to go to a public hospital and got my dialysis there. […] The cost of acquiring my dialysis medicines almost doubled, as they were not available at my regular pharmacist, my hospital was closed, I had to look everywhere and they were available in limited quantities only.”*

*“I have been advised to get three dialysis visits per week. Now because of the lockdown, I have only been able to get two per week. Hence, water fills in my lungs and I have difficulty breathing if I do any physical activity.”*

Family member of dead patient: *“We live in [District A] and the hospital he regularly visited was in [District B]. When the lockdown was imposed and there was a curfew, he had to find a smaller, alternative facility for dialysis. We were asked to get permissions and multiple slips from govt officials. He didn't get the care he usually received. That smaller hospital didn't do his dialysis properly and that's why he died. […] The medicines that were prescribed to him were only available in [District B] and not his local area due to the lockdown. He missed his medicines for BP and diabetes for 20 days.”*

Family member of dead patient: *“Due to the lockdown, the patient couldn't travel to [District B] on time and wasn't able to find a hospital in the vicinity, so he died on the way to the hospital itself.”*

### **Ethical considerations**

The research protocol and survey instruments were approved by the Institutional Review Boards of the Institute for Financial Management and Research (IFMR) in India and Stanford University in the United States. All data collection was managed by a team employed by JPAL South Asia at IFMR, which has extensive research conducting field and phone-based research. Standard research protocols were followed. Participants were informed of the nature and possible consequences of the study and were given the opportunity to discontinue participation at any point of their choosing.

Given that data were being collected during a pandemic that may have severely adversely affected households, the households in our sample may have been particularly vulnerable due to their socioeconomic status and illness, and the health and security of our data team was also a concern, we took several additional precautions to reduce any adverse consequences. To reduce the burden on households, we designed the surveys to be short: among surveyed households, the first round of the survey took 25 minutes and the second follow-up round took 13 minutes, on average. We trained surveyors to very clearly offer households the option not to participate, to be sensitive to households during the survey, particularly those that had faced substantial difficulties or deaths, and to provide all households with information on local COVID-19 and hospital health service helplines to consult if they experience medical problems. To protect the surveyors, all surveys were conducted over the phone from the security of their homes. Additional software was installed on the surveyor tablets to mask phone numbers from them and to automatically upload all data to secure cloud services without storing it locally.
